# Patient Perspectives on Home-Based Rehabilitation Exercise and General Physical Activity after Total Hip Arthroplasty: A qualitative study (PHETHAS-2)

**DOI:** 10.1101/2020.02.24.20025056

**Authors:** Anne Grøndahl Poulsen, Janni Dahlgaard Gravesen, Merete Nørgaard Madsen, Lone Ramer Mikkelsen, Thomas Bandholm, Camilla Blach Rossen

## Abstract

**Objectives:** To investigate patient-perceived facilitators and barriers to home-based rehabilitation exercise and general physical activity after THA.

**Design:** Using a qualitative design, twenty-two semi-structured interviews were conducted and analyzed using an interpretive thematic analysis approach, with theoretical underpinning from the concept conduct of everyday life. The study is embedded within the PHETHAS-1 trial, quantitatively investigating recovery outcomes after a home-based rehabilitation exercise program.

**Setting:** A regional hospital in Denmark between January 2018 and May 2019.

**Participants:** Twenty-two patients who had undergone THA and performed home-based rehabilitation exercise.

**Results:** The main theme “Wishing to return to the well-known everyday life” and the subtheme “General physical activity versus rehabilitation exercise” were identified. Generally, the participants found the home-based rehabilitation exercise boring but were motivated by the goal of returning to their habitual conduct of everyday life and perform their usual general physical activities. Participants enrolled in the PHETHAS-1 study used the enrollment as part of their motivation for doing the exercises.

Both pain and no pain were identified as barriers for doing the home-based rehabilitation exercise. Pain could cause insecurity while no pain could cause the rehabilitation exercise to be perceived as pointless.

**Conclusions:** The overall goal for the THA patients was to return to their habitual everyday life. This goal served as a facilitator for undertaking home-based rehabilitation exercise. Being able to perform their usual activities paradoxically became a barrier for some of the participants, as they were more motivated towards general physical activity than the rehabilitation exercise.

**Contribution of paper:** *Key messages:*

- Patients undergoing total hip arthroplasty have goal of returning to their habitual everyday life doing their usual physical activities.
- Home-based rehabilitation exercise can be perceived as boring and time-consuming and the goal of returning to their habitual everyday life serves as a facilitator to perform the rehabilitation exercise.

*What this paper adds:*

- Both pain and no pain can be a barrier to performing home-based rehabilitation exercise.
- Pain can cause uncertainty as to whether performing an exercise could be harmful, while no pain can cause the rehabilitation exercise to be perceived as pointless.
- Standard care patients tend to modify the rehabilitation program as they are gradually able to perform their usual general physical activities.
- Enrolment in clinical studies and contact with health professionals can facilitate adherence to home-based rehabilitation exercise.

**Trial identifier:** NCT03109821

## Introduction

Total hip arthroplasty (THA) is a common surgical intervention in western countries often performed as fast track surgery and the number of THAs has been rising [1,2]. Additionally, fast-track surgery has proven effective in terms of reducing costs, length of hospital stay, morbidity and convalescence [3, 4]. In Denmark alone 11.000 THAs are performed every year [5] with patients routinely discharged from the hospital within two days of surgery [6].

Rehabilitation exercise is custom as part of the post-operative program for patients undergoing THA in the expectation of reducing pain and increasing mobility [7]. This is also the case in Denmark, with each hospital having different procedures. Some hospitals refer patients to supervised rehabilitation exercise in the municipality while others use home-based rehabilitation exercise after an initial instruction [8].

Level 1a-evidence from systematic reviews show that supervised exercise after THA provides no additional benefit compared to home-based rehabilitation exercise after initial instruction with regard to patient-reported function, pain, health-related quality of life or performance-based functions [9,10]. Additionally, home-based rehabilitation is presumably less expensive than supervised rehabilitation and with rising health care costs, one might expect home-based rehabilitation exercise to become even more prevalent in the future.

There are indications that adherence to home-based rehabilitation exercise might be problematic [11], yet we know little about patients’ perspectives on home-based rehabilitation exercise and general physical activity after THA.

To be able to support and optimize clinical pathways with patients rehabilitating at home after THA, the PHETHAS studies were founded. PHETHAS-1 quantitatively investigates physical outcome of performing a home-based rehabilitation exercise program, while this study, PHETHAS-2, qualitatively investigates patient-perceived facilitators and barriers to home-based rehabilitation exercise and general physical activity after THA [12].

## Methods

### Theoretical underpinning

The concept conduct of everyday life from critical psychology [13,14] was chosen as a theoretical underpinning for this study. Conduct of everyday life is an overall concept that embraces the complexity of an individual’s everyday life across contexts [13,14]. It includes the different aspects of a person’s everyday life which could be working, performing general physical activities or home-based rehabilitation exercises. According to theory the individual person prioritizes activities based on what he/she considers as contributing to their subjective understanding of quality of life [13,14].

Using conduct of everyday life as theoretical underpinning provides the potential to elucidate how patients integrate both general physical activity and home-based rehabilitation exercise into their everyday lives in the rehabilitation period, and thereby enlighten possible patient perceived barriers and facilitators for performing the rehabilitation exercise.

We defined home-based rehabilitation exercise as a plan of physical activities designed and prescribed to meet specific therapeutic goals. Its purpose is to restore normal musculoskeletal function or to reduce pain caused by diseases or injuries. This definition is synonymous with the MeSH term “Exercise therapy” as defined in the PubMed MeSH database [15] and in alignment with WHO’s description of ‘exercises’ as a subcategory of ‘physical activity’ [16]. In this study we distinguish this type of prescribed rehabilitation exercise from general physical activity undertaken while working, playing, gardening and engaging in leisure time activities.

### Participants and recruitment

Participants were recruited from a Danish Regional Hospital in the period January 2018 to May 2019. Inclusion criteria were adults > 18 years who underwent a primary THA due to osteoarthritis and exclusion criterion was referral to supervised rehabilitation. The participants should be able to understand written and spoken Danish. The participants were purposely sampled [17] aiming to reflect gender and age in the group of typical THA patients [18]. Twenty-two participants were included. The demographics of the participants are illustrated in table 1.

**Table 1.** Characteristics of participants

|  | Participants recruited from<br>standard care<br>(n=8) | Participants recruited from<br>PHETHAS-1*<br>(n=14) | All<br>participants<br>(n=22) |
| --- | --- | --- | --- |
| Gender<br>(female/male)<br>Number (percent) | 6/2 (75/25) | 4/10 (29/71) | 10/12 (45/55) |
| Age (years)<br>Median (range) | 70 (42-82) | 69 (48-80) | 69 (42-82) |
| Retired (yes/no)<br>Number (percent) | 5/3 (63/37) | 10/4 (71/29) | 15/7 (68/32) |

\* Pragmatic Home-based Exercise after Total Hip Arthroplasty – Silkeborg (PHETHAS-1) is a trial investigating the preliminary efficacy of home-based rehabilitation using elastic band exercise on performance-based function after THA.

As this study was embedded in the PHETHAS-1 study, participants were recruited from participants enrolled in PHETHAS-1 by the researcher responsible for PHETHAS-1 (MNM). To avoid gathering data only from participants in PHETHAS-1, who might be more motivated for exercise than the average THA-patient and hence more adherent than those who decline participation in clinical exercise trials, we recruited additional eight participants from standard care. Standard care participants were recruited by physiotherapists responsible for the standard care pathway at the hospital. (See illustration 1: flowchart of pathways for both standard care and PHETHAS-1 participants). All participants were instructed in performing the exact same home-based rehabilitation exercise. Details of this home-based rehabilitation exercise have previously been published [10], see illustration 1 for an overview.

Insert Illustration 1 here: flowchart of participant pathways, recruitment and content of the rehabilitation program.

### Data collection

Individual interviews with the participants were conducted to gain in-depth knowledge about experiences with home-based rehabilitation exercise and general physical activity after THA [19]. The interviews were guided by a semi-structured interview guide [19, 20] (Appendix A), which was informed by existing knowledge in the field of THA along with the theoretical concept conduct of everyday life. Data collection and sampling was guided by a concurrent data analysis [21]. The interviews were conducted 10 weeks postoperatively. The interviewer obtained informed consent from the participant prior to the interview.

Participants enrolled in PHETHAS-1 were physically tested at the hospital 10 weeks postoperatively and were interviewed afterwards in an undisturbed meeting room. Participants following standard care were interviewed in their homes (illustration 1). Interviews were audio recorded and lasted an average of 43 minutes (20–67 minutes). The interviews were conducted by AG, JG and CR. AG and JG both hold a master’s degree, are trained in the qualitative field of research and are trained interviewers. They were supervised by CR, who is an experienced qualitative researcher and interviewer.

The study complies with the declaration of Helsinki [34], was approved by The Ethics Committee of Central Denmark Region and the Danish Data Protection Agency (ref. no: 1-16-02-589-15).

### Data analysis

Data were thematically analyzed which is a method for identifying and analyzing patterns and themes across data [22,23]. The audio recordings were transcribed vertibam by assistants and listened, read and corrected by AG and JG. Initially, the interviews were manually coded by AG and JG, supervised by and discussed with CR. The coding process was guided by how the participants integrate home-based rehabilitation exercise and general physical activity as part of their everyday life during the rehabilitation period, e.g. which activities they prioritized and why, along with factors serving as motivators or barriers respectively, but also with an openness for other themes of importance. After potential themes were identified and discussed with co-authors, original data were re-visited to validate themes within the data-set in an iterative process [22,23]. The analytic process was supported by NVivo 12 software [24].

## Results

The analysis resulted in the main theme “Wishing to return to the well-known everyday life” which is elaborated on below including the subtheme “General physical activity versus rehabilitation exercise”.

### Wishing to return to the well-known everyday life

All participants wished to be physically active and their overall goal was to return to their habitual everyday life. The following quote came from a male participant who was still working and enjoyed being active with sports.

> *P04: The goal was to be able to do sports again. Primarily to be pain free. And then leading a more or less ordinary life again with some sports. [*…*] Aesthetically it is nice to get outside and experience the world with your eyes and ears, as you do when you go outside. First and foremost, the aim of the operation was to get my quality of life back again and then pain free*.

The participants’ goal was to return to their usual everyday life consisting of activities that contribute to their quality of life. What they perceived as valuable activities were individual, and they used their own habitual everyday life activities as a reference point. Participants found motivation for doing the home-based rehabilitation exercise program in the belief that it would bring them closer to their goal.

When asked whether there were times when it was difficult to get the home-based rehabilitation exercise done, a male participant working part time in a shop answered:

> *P07*: *No. If something came up, I did it in the evening*.[…*] I think it is nice you can decide for yourself when you do it, compared to going somewhere to see a physiotherapist*.

Analysis showed that flexibility on when and where to include the rehabilitation exercise into their everyday life facilitated performance of the rehabilitation exercises for some participants. The flexibility made room for other activities they considered contributing to their quality of life. In that sense the home-based rehabilitation exercises would have an advantage compared to supervised rehabilitation.

A few participants missed being in contact with a physiotherapist during the rehabilitation period. A participant retired from an office job, described how she phoned the hospital staff to address certain issues she worried about. She would prefer participating in group training with a physiotherapist.

> *P12: Because first of all you could have your exercises corrected. Second you could have been told when to use a tighter elastic band. And talk to the others. And this thing in my head being so afraid of crossing my legs, I think that could have been killed [laughing]. And then I might have been able to talk about pain in the groin, because I did have a lot of pain in the groin. Just being told to go see my own [private] physiotherapist with that problem. That would have been nice*.

Lack of contact with a physiotherapist during the period of performing the home-based rehabilitation exercise could be identified as a possible barrier for some participants. The quote above reflects missing the possibility to address issues of uncertainty with both a physiotherapist and other THA patients.

Our data show that most participants experience a degree of pain that did not affect their performance of the home-based rehabilitation exercise. But analysis revealed that both more intense pain and no pain could affect performance of home-based rehabilitation exercise. A male participant usually being active and working in an academic job experienced rather intense pain and described his struggle.

> *P06: I think the challenge all along has been how much it must hurt. We are instructed to repeat to exhaustion,[…]but where is that point when you are in pain?*

Other participants felt hardly any pain at all. An active female participant, who was retired from the healthcare sector, explained how having no pain affected her.

> *P14: When you get out of bed 3 hours after the surgery and walk and bike and climb stairs and go all the way down the hall and back again and you notice nothing. Then you say to yourself: nothing is wrong with you*.*[…] Then you really have to pull yourself together to do the exercises, because you already feel that you can do everything*.

Our data paradoxically showed that both pain and no pain can be seen as barriers in regards to performing the home-based rehabilitation exercise. Pain provided insecurity about how to best perform the exercise and no pain provided the possibility of simply returning to the habitual and preferred everyday life, which was more tempting than prioritizing time to do the rehabilitation exercise.

### General physical activity versus rehabilitation exercise

The subtheme ‘General physical activity versus rehabilitation exercise’ showed that the participants consistently distinguished between the instructed home-based rehabilitation exercise and general physical activities they considered part of their habitual everyday life.

This retired female participant was usually very active with hiking and fitness. She explained:

> *P001: Well, I would rather do normal activities, long walks or something like that. That’s what I prefer. And I do the exercises to achieve that. I mean, it is quite boring doing those exercises, it depends on what’s on the radio [laughs]*.[…*]I do them to be able to do the other things*.

For most participants, the rehabilitation exercise was used as a means of regaining their habitual everyday life. Their usual general physical activities contributed to their understanding of quality of life and were considered joyful, while the rehabilitation exercise was perceived as boring and time consuming.

A male participant had already started work and fitness and explained why he no longer performed the rehabilitation exercise as instructed.

> *P008: It is going so well [laughing]. I do them, [the exercises] but not*…*maybe not every day, and there are days where I have forgotten*.

Analysis revealed a difference between the group of standard care participants and participants also enrolled in PHETHAS-1. Standard care participants often modified the home-based rehabilitation exercises as illustrated in the citation above. To this group the rehabilitation exercises were perceived as losing their purpose, as their level of functioning improved. and they were able to perform usual general physical activities.

In contrast a very active male participant enrolled in PHETHAS-1 explained his motivation for performing the exercises.

> *Interviewer: [*…*]As you have resumed all these usual activities, are you still motivated for doing the exercises with the elastic band?*
>
> *P08: I think so, yes. Absolutely, because it is part of this trial [PHETHAS-1] that I wish to be very loyal to. So I have followed it completely. Otherwise you can’t use it for anything if you don’t know whether one just filled it [the training diary] out as one pleases*.

Analysis showed that for the group of participants enrolled in PHETHAS-1, their enrollment served as a facilitator for performing the rehabilitation exercises exactly as instructed. They referred to an obligation towards the researcher and the study they had signed up for, also knowing that it would supposedly benefit themselves as well.

## Discussion

Findings from our study show that participants wished to return to their habitual conduct of everyday life after their THA surgery. Similar results have previously been reported in other studies [25, 26] also concluding that patients have little interest in achieving greater levels of physical activity, than they had before the hip restricted their functioning [25]. In our study, many participants found the home-based rehabilitation exercise boring and preferred performing usual general physical activities. They generally performed the home-based rehabilitation exercise, believing it would bring them closer to their goal of returning to their usual conduct of everyday life and used this as a motivator to get the exercise done.

Furthermore, we found an important difference between the two groups of participants. Analysis showed that participants also enrolled in PHETHAS-1 were motivated by an obligation towards the study and the researcher, which support a review with similar findings [27]. Other studies also find that contact with a physiotherapist can enhance adherence to rehabilitation exercises, using the concept of therapeutic alliance as a possible explanation [28,29]. Therapeutic alliance focuses on the impact of the relation between patient and professional on adherence to rehabilitation exercise [28,29]. This knowledge is crucial when assessing results from clinical training studies.

Standard care participants gradually modified the exercises, when they were able to return to their habitual everyday life, performing usual general physical activities they felt contributed to their quality of life. Modifying therapeutic instructions is well known in other areas [30], but to our knowledge, it is the first time to be described in THA-patients.

The participants generally favored general physical activities when possible, and it would be relevant to investigate whether general physical activities could be as effective as home-based rehabilitation exercise and if so, whether future THA-patients could rehabilitate doing only their preferred physical activities.

A previous study showed that patients can have a feeling of being left alone after discharge from the hospital, for example in dealing with pain [31]. Our study support this finding along with participants describing the wish of having contact to a physiotherapist who can give advice. Furthermore our study adds the knowledge that no pain also can be identified as a barrier for performing home-based rehabilitation exercise.

### Strengths and limitations

While it could be seen as a limitation of the study that we recruited participants from both the PHETHAS-1 study and from standard care, this particular combination of participants revealed an important difference in motivation towards adherence to, and performance of, the home-based rehabilitation program and could therefore be viewed as a strength. Moreover, we note that our findings show no other differences between the two groups of participants.

We recruited participants from only one hospital and since the rehabilitation after THA differs between hospitals this might have affected our results, although we made clear what this particular rehabilitation consisted of.

The participants in our study are considered relatively physically active, which could have influenced the results and it would have been desirable to have included more sedentary patients as well. Also our participants consisted of fewer females compared to the general group of patients undergoing a THA [18], which might have affected our results.

There may be additional contributing factors in relation to patients’ perceptions of home-based rehabilitation exercise after THA such as age, gender, previous training experience and culture. Further studies are needed to explore this.

Scientific rigor is enhanced by using theory throughout the scientific process [32] and triangulation in form of more investigators collaborating on the analysis, is also considered a strength [33].

## Conclusion

This study showed that THA patients’ goal of returning to their habitual conduct of everyday life as it was before their hip restricted their functioning generally served as a facilitator for performing the home-based rehabilitation exercise, because it was perceived as a means to achieve their goal. Most participants found the rehabilitation exercises boring and would prefer usual general physical activities. Partly motivated by an obligation towards the researcher, participants enrolled in the PHETHAS-1 trial reportedly performed the home-based rehabilitation program as instructed. In contrast, the participants following standard care often modified the program, as they got able to perform their usual general physical activities, which they found more motivating.

Paradoxically both pain and no pain could be identified as barriers for performing the home-based rehabilitation exercise and some participants experiencing barriers towards the home-based rehabilitation exercise missed having contact to a physiotherapist in the rehabilitation period.

## Data Availability

The data that support the findings of this study are available on request from the corresponding author. The data are not publicly available due to  the privacy of research participants.

Illustration 1: Flowchart of the study design in PHETHAS-2.

See separate file.

## Appendix 1.

### Interview guide

#### Introduction

- Tell a little about yourself: education, work life, leisure time, marital status

The interviewer presents the Timeline (a piece of paper with the illustration below)

**Surgery 3 week visit 10-week visit**

#### Questions asked while using the timeline

- What did your hip problems prevent you from doing?
- On a scale from 0 to 10, where 10 is best, how was your functional status just before the operation?
- Why was it like that?
- Where are you on that scale now?
- Where you nervous before the operation?
- Do you know anyone who has undergone hip surgery?
- What is your goal?

#### Questions concerning exercise and physical activity

- Which recommendations have you received concerning training from the physiotherapist (staff)?
- Tell me about how it was for you to perform the training at home just after discharge from the hospital?
- How was it to perform the training at home after the 3 week visit at the Hospital?
- Did you feel sufficiently informed/capable to independently conduct the training at home?
- Tell me how you do in practice when you exercise at home?
- Besides from the prescribed exercises, do you perform any other physical activity during the day? (e.g. biking, walking). (Explore patient preferences for physical activity versus exercise)
- How did it feel to “exercise to exhaustion”?
- What do you think about the exercise diary?
- Have you had periods where it was difficult to exercise/where you did not perform exercises?
- How important is it for you to be able to be physically active?
- Have you considered not to be physically active/perform the exercises?
- Have you considered when you will quit the exercise program?
- Have you considered if you wish to continue to exercise and how you will do it?

#### Supplementary question

We are considering to investigate whether this exercise program works better than general physical activities, what do you think of this

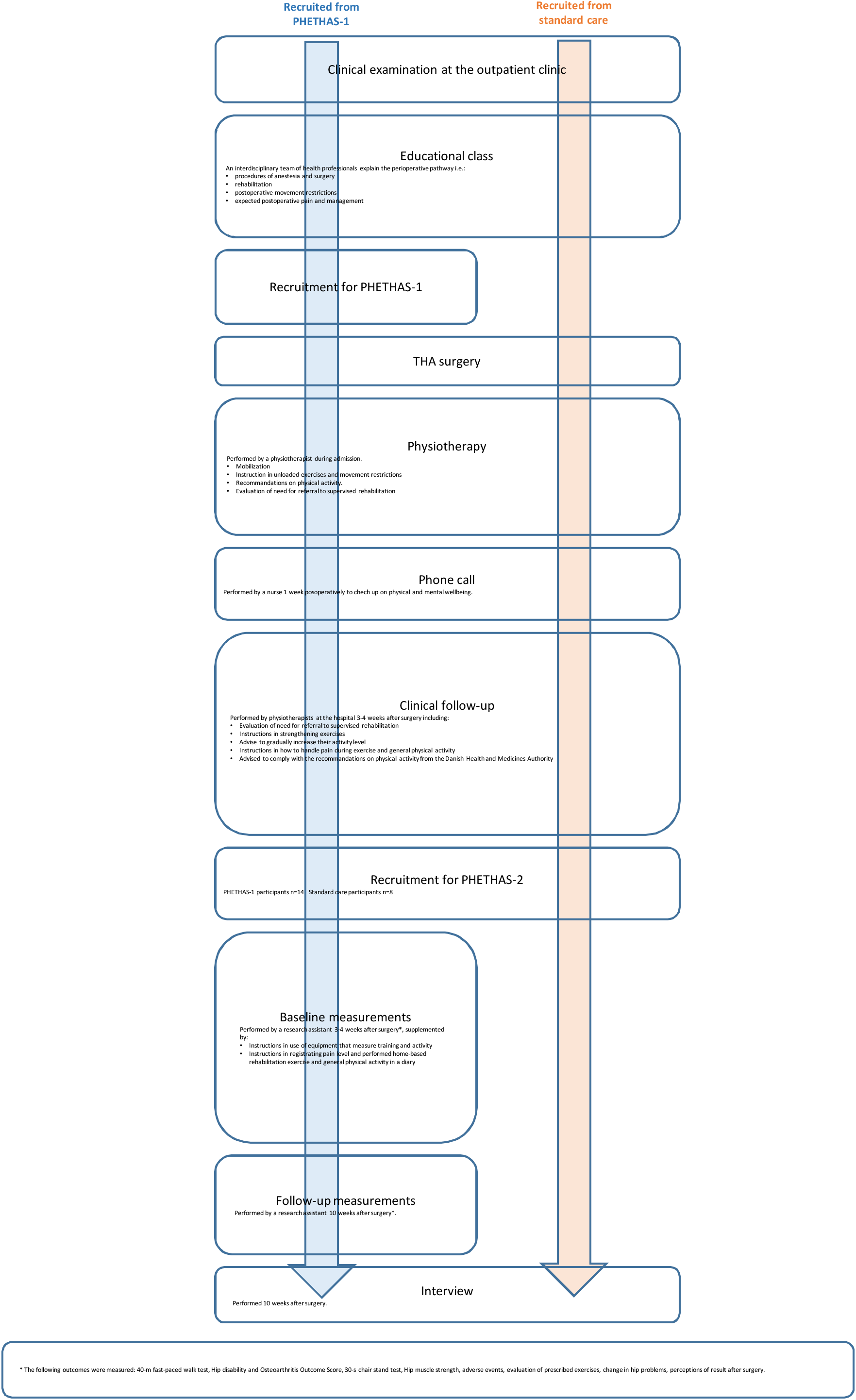

